# The illicit cigarette market in the Democratic Republic of the Congo (DRC): findings from a cross-sectional study of empty cigarette packs

**DOI:** 10.1101/2024.10.25.24316163

**Authors:** Noreen Dadirai Mdege, Christelle Tchoupe, Retselisitsoe Pokothoane, Didier Mirindi, Christus Cito Miderho, Kelley Sams, Patrick Shamba, Emmanuel Kandate, Patrice Milambo, Hana Ross

**Author notes:** Corresponding author: (NDM).

## Abstract

This study aimed to estimate the proportion of cigarettes consumed in the Democratic Republic of the Congo (DRC) that are illicit and the extent of cigarette tax evasion; and to identify the origins of and factors associated with illicit cigarettes. Data were collected from May 15 to June 9, 2023. Stratified, multistage sampling was used to select 32 health areas from which empty cigarette packs were collected. Each collected pack was examined and classified as licit if it complied, or illicit if it did not comply, with the DRC’s tax stamp or written health warning requirements, or the requirements to have a notice indicating the prohibition of sale by/to minors or information on tar and nicotine content. We reported frequencies as numbers and percentages, and means for continuous variables. We performed regression analysis and used adjusted odds ratios (aOR) and their 95% confidence intervals (CI) to measure associations. 8.6% of the 10622 empty cigarette packs collected were illicit. All of the illicit packs were also identified as having evaded cigarette tax. Approximately 8.0% of the collected packs did not comply with written health warning requirements, 5.6% did not indicate the prohibition of sale by/to minors, and 4.5% did not have information on tar and nicotine content. Packs from low-income areas were more likely to be illicit than those collected from high-income areas (aOR 1.90; [95% CI: 1.48-2.43]). The likelihood of being an illicit cigarette increased with increasing susceptibility to armed conflict/insecurity. Packs from border provinces were less likely to be illicit than those from non-border provinces (aOR 0.48; [95% CI: 0.25-0.90]). Our results show that the illicit cigarette trade market in the DRC is currently small, and most illicit cigarettes are imported from other countries. There is a need to secure the cigarette supply chain, including strengthening border controls.

## Background

In 2017-2018, 18% of men and 2% of women aged 15 to 49 years used tobacco in the Democratic Republic of the Congo (DRC), and 13% and <1% smoked cigarettes, respectively [1]. The DRC has progressively developed and implemented regulatory measures aimed at reducing the consumption of tobacco products over the past three decades. These include prohibiting the advertising, promotion and sponsorship of tobacco, tobacco products and their derivatives; banning smoking in public places; mandating written health warnings on tobacco products; and tobacco taxation, although this is at 38.7% of the retail price, and falls short of the recommended 75% [2].

In 2004, the country signed the World Health Organization Framework Convention on Tobacco Control (WHO FCTC) which provides evidence-based policy interventions to address the tobacco epidemic and its consequences, and ratified the treaty in 2005 [3]. In addition, the country also signed the Protocol to Eliminate Illicit Trade in Tobacco Products in 2013 in recognition of the importance of combating illicit trade in tobacco products by securing the supply chain, although this is yet to be ratified [4]. The illicit trade of tobacco products undermines tobacco control and public health by increasing access to the products, often cheaper ones, and thereby increasing consumption [5–8]. The Protocol to Eliminate Illicit Trade in Tobacco Products covers measures such as track and trace systems, licensing, due diligence, and issues related to internet- and telecommunication-based sales, tobacco product transactions in free zones and international transit, and duty-free sales [9].

The DRC shares its borders with nine countries: the Republic of Congo, Uganda, Burundi, Rwanda, Tanzania, the Central African Republic, Sudan, Zambia and Angola. The DRC has experienced persistent social and political instability throughout its recent history which contributes to porous borders and increases its susceptibility to the illegal importation of cigarettes from neighbouring countries. However, the extent of illicit cigarette trade in the DRC has not been investigated before, so its size, trend and characteristics are not documented or well understood. This hampers tobacco control decision making, and is particularly important because the tobacco industry often uses illicit tobacco trade as a reason to oppose effective tobacco control measures such as tax increases [10]. This study, therefore, aimed to 1) estimate the proportion of cigarettes consumed that are illicit, 2) estimate the extent of cigarette tax evasion, 3) identify the origins of illicit cigarettes, and 4) identify factors associated with illicit cigarettes.

## Methods

### Study design

We conducted a national-level, cross-sectional study in which empty cigarette packs were collected [11, 12] from three types of collection points: stationary retailers, mobile retailers, and garbage bins/streets. Stationary retailers sell from the same location (e.g., shops, kiosks, and stalls), while mobile retailers are itinerant and sell cigarettes on the move. By law, the sale of single cigarette sticks is not prohibited in the DRC and is highly prevalent, which makes it possible to collect empty cigarette packs from retailers. We also collected average cigarette retail price information for each brand from retailers.

### Study setting

The sampling of study locations was based on the structure of the DRC health pyramid which spans from provinces, to health zones (HZs) within provinces, to health areas (HAs) within HZs. We stratified the 26 provinces of the DRC into four groups based on the level of porosity, i.e., border/non-border status and susceptibility to armed conflict/insecurity. Stratum 1 had two provinces with very high-porosity (i.e., at the border and highly susceptible to armed conflict); stratum 2 had one province with high-porosity (at the border and the capital); stratum 3 included 14 provinces with intermediate-porosity (i.e., border provinces that are not subject to armed conflicts); and stratum 4 consisted of nine provinces with low-porosity (i.e., non-border provinces with no particular security issues) (S1 Fig). We randomly sampled provinces from each stratum with the number of provinces to be drawn from each stratum being proportional to the weight of the stratum. The total number of provinces included in the study was eight: one province each from stratum 1 (Ituri) and 2 (Kinshasa), four provinces from stratum 3 (Haut-Katanga, Kasaï-Central, Kwango, Nord-Ubangui), and two provinces from stratum 4 (Tshopo and Sankuru).

For each sampled province, we grouped the HZs into two strata, i.e., urban and rural, and we randomly sampled one HZ from each of the strata. The third level of sampling involved randomly sampling two HAs from each participating HZ (S2 Fig). This yielded a total of 32 HAs as sites for data collection (S1 Table).

### Criteria for identifying illicit empty cigarette packs

By law, there are a number of requirements regarding what should be on a cigarette pack sold in the DRC. These include a notice on the ban on the sale of tobacco to minors and by minors, written health warnings, brand identifiers, information on tar and nicotine content, and an affixed tax stamp. In addition, misleading names, words or logos, for example, those that are likely to create confusion or give the impression that a particular brand can promote fitness and well-being in general, are prohibited. We conducted semi-structured one-on-one discussions and workshops with key tobacco control stakeholders in the DRC, including the Programme National de Lutte Contre la Toxicomanie et les Substance Toxiques (National Program for the Control of Drug Addiction and Toxic Substances), Direction Générale des Douanes & Accises (Directorate-General of Customs and Excise), and the Société Industrielle et Commerciale des Produits Alimentaires (Industrial and Commercial Company of Food Products), to agree on the definition of an illicit cigarette pack. From consensus with these tobacco control stakeholders, an empty cigarette pack was classified as illicit if it met any of the following criteria:

● *cigarette tax evasion/non-compliance with tax stamp requirements* (i.e., no tax stamp, has a duty-free stamp but collected from a retailer who is unauthorised to sell duty-free cigarettes, or has the wrong stamp). Every cigarette pack in the DRC should have a tax stamp that is orange if for locally manufactured cigarettes that are intended for domestic consumption, grey for imported cigarettes, or green for cigarettes intended for duty-free shops.
● *non-compliance with written health warning requirements* (no written health warnings on the two main sides, or has written health warnings in a language other than French). Every pack of cigarettes must bear at least two of the following four health warnings in French: “Smoking is harmful to health”, “Tobacco seriously harms your health”, “Be careful, smoking kills”, or “Smoking is highly addictive”.
● *absence of the notice indicating the prohibition of sale by/to minors*. By law, each pack of cigarettes must bear the words “Prohibited for sale to minors and by minors.” This notice must be printed in bold, indelible, and visible capital letters, with a height of at least two millimetres, on the top of the right side of the pack.
● *no information on tar and nicotine content.* Tar and nicotine content information should be on the right side of each pack, and cover at least 20% of the side.

During data collection, we found that some empty cigarette packs had a yellow tax stamp. This is an old stamp that was used for both imported and locally manufactured cigarettes, and according to the Direction Générale des Douanes & Accises, cigarettes imported after a transitional period of 60 days from April 15, 2022 should not bear this stamp. However, there is a lack of clarity on whether those manufactured in the DRC, or those imported before the transition period but are still on the shelves in the DRC can still have the yellow stamp. In addition, there was recognition from some key tobacco control stakeholders that the transition from yellow stamps is not yet complete.

### Data collection

Data were collected from May 15 to June 9, 2023. We aimed to collect at least 10,000 empty cigarette packs in total from the eight provinces. This quantity was based on information from similar studies [12, 13]. Data were collected by a team of 40 data collectors and eight supervisors. The team was trained in the study protocol and data collection process, including the recruitment of retailers, and the collection of empty cigarette packs from retailers and garbage bins/streets. The team was also trained on the use of data collection instruments, in particular, the questionnaires that were programmed on tablets, and the Distance Meter application for measuring distance to ensure that the data were collected only from the demarcated data collection grid. Considering the size of the DRC, training took place in two phases: 1) the training of the eight supervisors (one supervisor per province) from April 3 to 6, 2023; and 2) the training of data collectors by the supervisors from April 10 to May 14, 2023. Pilot tests were carried out in each province after the completion of classroom training.

### Empty cigarette pack collection

Data collectors identified the centre of economic activity in an HA (e.g., the main market) to serve as the starting point for data collection. They then collected empty cigarette packs from consenting retailers and garbage bins/streets within a ∼100m x 100m square grid (in urban areas) or a circle grid of ∼800m in diameter (in rural areas) around the starting point [14]. For the collection of packs from retailers, data collectors approached retailers within the grid and informed them about the study. Those who were interested in study participation were provided with written study information and requested to provide verbal consent if they agreed to participate. Consenting retailers were supplied with a prelabelled bag the same day and asked to deposit all empty cigarette packs from single stick sales in the bag. The bags with empty packs were retrieved the following day. Each collection bag had a unique identifier to distinguish between retailers, HAs, HZs, provinces, and types of collection point (i.e., stationary retailers, mobile retailers, and garbage bins/streets).

### Data extraction from empty cigarette packs

Data were recorded using a questionnaire programmed in SurveyCTO [15]. We recorded area-level information including the province, porosity stratum, HZ, HA, rural/urban, and area-level income group. Retailer-level information included the type of retailer and total number of packs collected (including by brand). For each empty cigarette pack, we extracted information that included the brand name, manufacturer, and country of origin. We also recorded whether the pack had a tax stamp, whether the tax stamp was a DRC mandated stamp, and the colour of the stamp. We recorded whether the pack had a written health warning, how many, the position and wording of each warning, the language; whether the health warnings were printed in bold capital letters, with black on a contrasting white background, and covered 30% of the two main presentation areas of the pack; and whether the background reserved for health warnings was framed by a distinctive black outline printed in bold. The collected information also included whether the nicotine and tar content were stated, the location of this information on the pack and whether it covered 20% of the side on which it was located. We also collected information on whether the pack had the phrase “PROHIBITED FOR SALE TO AND BY MINORS”; whether this was printed in bold capital letters, indelible and visible, and was at least 2 mm high on the top right side of the pack.

### Quantitative survey with retailers

We used an interviewer-administered questionnaire to collect cigarette retail price data from those retailers provided the empty cigarette packs. The questionnaire also collected information on where retailers source their cigarettes, the brands of cigarettes they sell, the sale and purchase prices of cigarettes, and the socio-demographic characteristics of the retailers. The questionnaire was also programmed in SurveyCTO [15].

The questionnaires used for this study were informed by literature reviews and guidelines/toolkits for measuring the illicit trade of tobacco products [14, 16]. The questionnaires were pretested before use and revised for clarity and relevance.

### Data analysis

Our primary analysis considered packs with yellow stamps as legal if they did not meet any other ‘illicit’ criteria. We conducted descriptive statistics to estimate the overall proportion of empty cigarette packs (the proportion of cigarettes consumed) that was illicit, as well as by type of collection point, province, porosity stratum, rural/urban, area-level income group, imported/locally manufactured, brand and country of origin. Additionally, the proportion of illicit packs was calculated based on each of the specific criteria, i.e., cigarette tax evasion/ non-compliance with tax stamp requirements, non-compliance with written health warning requirements, absence of the notice indicating the prohibition of sale by/to minors and no information on tar and nicotine content. The extent of tax evasion was determined by focusing on the proportion of cigarette packs that did not have a tax stamp, had the wrong tax stamp or had a duty-free stamp but were obtained from a retailer that is not authorised to sell duty-free cigarettes. The frequencies were reported as numbers and percentages. We also calculated the average retail price for a cigarette pack by brand.

We used logistic regression to examine the factors associated with being an illicit cigarette pack, using adjusted odds ratios (aOR) and their 95% confidence intervals (CI) as measures of association. We considered the following independent variables based on literature that suggest their association with the prevalence of illicit cigarette consumption [17, 18]: border/non-border province, area-level income status (low/high), urban/rural health zone, collection point (stationary retailer/mobile retailer/garbage bin or street), porosity stratum, country of origin, province, cigarette manufacturer, cigarette brand, and whether the cigarettes were flavoured or not. Due to high multicollinearity between some explanatory variables, we dropped three independent variables: country of origin and province which were strongly correlated with porosity stratum, and cigarette brand which was strongly correlated with country of origin. High- and very high-porosity strata were combined into one category in the regression analysis. Statistical significance was set at the 0.05 level. The analyses were performed using STATA Version 17.0 [19].

We conducted sensitivity descriptive analyses in which we considered all imported packs with a yellow stamp to be illicit.

### Ethics approval and consent to participate

Ethics approval was obtained from the National Health Ethics Committee of the Ministry of Health of the DRC on May 3, 2023 (approval number 443/CNES/BN/PMMF/2023). The participating retailers were provided with study information, including the study objectives, the procedures for participation, and the right to abstain from participation in the study or to withdraw consent to participate at any time without reprisals. They provided written informed consent to participate in the study before taking part.

## Results

### General characteristics of the empty cigarette packs

A total of 10,622 empty cigarette packs were collected, of which 67.0% (7,116) were from stationary retailers, 14.3% (1,522) were from mobile retailers and 18.7% (1,984) were from garbage bins/streets (Table 1). Kwango province had the highest number of packs collected (1,813; 17.1%) whilst the lowest number was from Sankuru province 583 (5.5%). Very high-porosity provinces contributed 13.9% (1,479) of the packs collected, whilst high-porosity provinces contributed 16.4% (1,740), intermediate-porosity provinces contributed 53.7% (5,702) and low-porosity provinces contributed 16.0% (1,701).

**Table 1.**
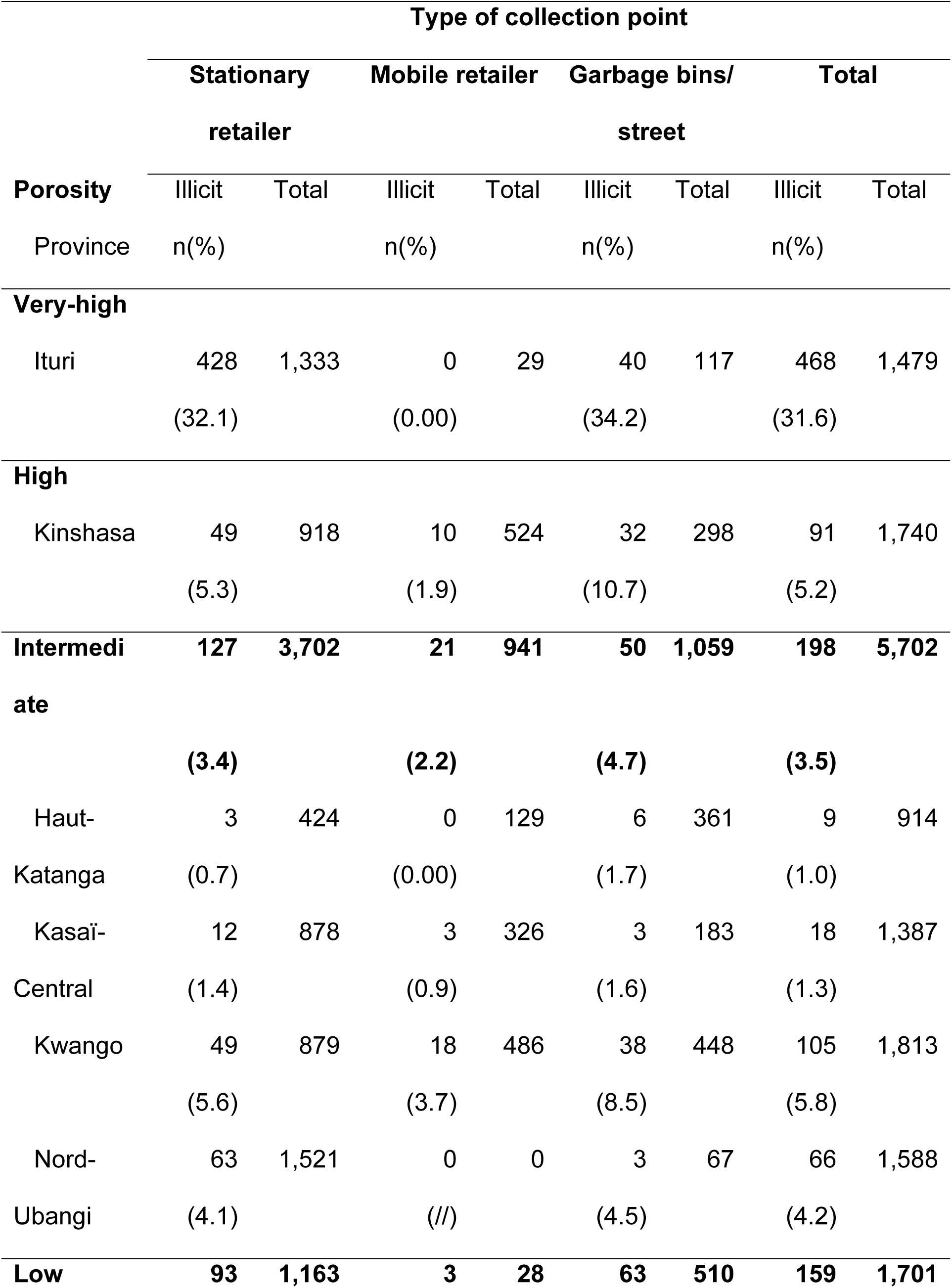
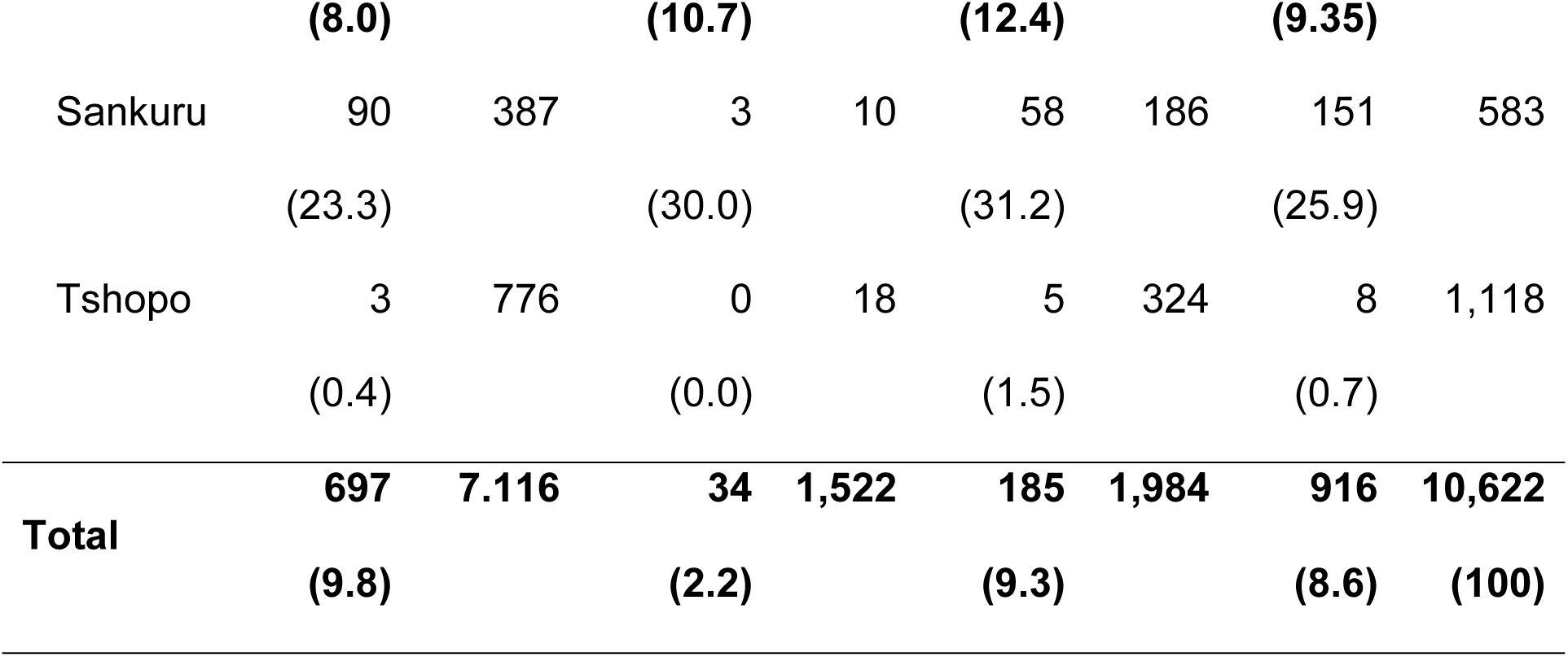
Proportion of illicit empty packs by type of collection point, porosity and province.

| **Porosity** | **Type of collection point** |  |  |  |  |  |  |  |
| --- | --- | --- | --- | --- | --- | --- | --- | --- |
|  | **Stationary** |  | **Mobile retailer** |  | **Garbage bins/** |  | **Total** |  |
|  | **retailer** |  |  |  | **street** |  |  |  |
|  | **Illicit** | **Total** | **Illicit** | **Total** | **Illicit** | **Total** | **Illicit** | **Total** |
| Province | n(%) |  | n(%) |  | n(%) |  | n(%) |  |
| **Very-high** |  |  |  |  |  |  |  |  |
| Ituri | 428 | 1,333 | 0 | 29 | 40 | 117 | 468 | 1,479 |
|  | (32.1) |  | (0.00) |  | (34.2) |  | (31.6) |  |
| **High** |  |  |  |  |  |  |  |  |
| Kinshasa | 49 | 918 | 10 | 524 | 32 | 298 | 91 | 1,740 |
|  | (5.3) |  | (1.9) |  | (10.7) |  | (5.2) |  |
| **Intermedi** | **127** | **3,702** | **21** | **941** | **50** | **1,059** | **198** | **5,702** |
| **ate** | **(3.4)** |  | **(2.2)** |  | **(4.7)** |  | **(3.5)** |  |
| Haut- | 3 | 424 | 0 | 129 | 6 | 361 | 9 | 914 |
| Katanga | (0.7) |  | (0.00) |  | (1.7) |  | (1.0) |  |
| Kasaï- | 12 | 878 | 3 | 326 | 3 | 183 | 18 | 1,387 |
| Central | (1.4) |  | (0.9) |  | (1.6) |  | (1.3) |  |
| Kwango | 49 | 879 | 18 | 486 | 38 | 448 | 105 | 1,813 |
|  | (5.6) |  | (3.7) |  | (8.5) |  | (5.8) |  |
| Nord- | 63 | 1,521 | 0 | 0 | 3 | 67 | 66 | 1,588 |
| Ubangi | (4.1) |  | (//) |  | (4.5) |  | (4.2) |  |
| **Low** | **93** | **1,163** | **3** | **28** | **63** | **510** | **159** | **1,701** |
|  | **(8.0)** |  | **(10.7)** |  | **(12.4)** |  | **(9.35)** |  |
| Sankuru | 90 | 387 | 3 | 10 | 58 | 186 | 151 | 583 |
|  | (23.3) |  | (30.0) |  | (31.2) |  | (25.9) |  |
| Tshopo | 3 | 776 | 0 | 18 | 5 | 324 | 8 | 1,118 |
|  | (0.4) |  | (0.0) |  | (1.5) |  | (0.7) |  |
| **Total** | **697** | **7,116** | **34** | **1,522** | **185** | **1,984** | **916** | **10,622** |
|  | **(9.8)** |  | **(2.2)** |  | **(9.3)** |  | **(8.6)** | **(100)** |

When considering urban/rural classification, this was 74.5% (7,918) and 25.5% (2,704) of the packs, respectively (Table 2). 67.1% (7,130) of the packs came from low-income areas and 32.9% (3,492) came from high-income areas. 45.5% (4,834) of the packs were of cigarettes manufactured in the DRC, and the rest were of imported cigarettes. Four brands, i.e., the Equateur, Monte Carlo, Master, and Pall Mall accounted for 66.6% of the packs collected. None of the collected cigarette packs had a duty-free stamp.

**Table 2.** Proportion of illicit empty packs by rural/urban, area-level income group, brand and country of origin.

|  | **Total packs**<br>**collected** | **Number of**<br>**illicit packs** | **Proportion of**<br>**illicit packs** |
| --- | --- | --- | --- |
| **Urban/rural** |  |  |  |
| Rural | 2,704 | 219 | **8.1** |
| Urban | 7,918 | 697 | **8.8** |
| **Area-level income group** |  |  |  |
| High-income | 3,492 | 357 | **10.2** |
| Low-income | 7,130 | 559 | **7.8** |
| **Brand** |  |  |  |
| Supermatch | 894 | 465 | **52.0** |
| Oris | 532 | 380 | **71.4** |
| Pall Mall | 1,238 | 16 | **1.3** |
| Monte Carlo | 1,914 | 14 | **0.7** |
| Stella | 546 | 8 | **1.5** |
| Business | 147 | 8 | **5.4** |
| Caesar | 120 | 1 | **0.8** |
| Ambassade | 240 | 1 | **0.4** |
| Elite | 525 | 0 | **0.0** |
| Equateur | 2109 | 0 | **0.0** |
| Master | 1809 | 0 | **0.0** |
| Portsman | 262 | 0 | **0.0** |
| Other brands | 286 | 23 | **8.0** |

| **Country of origin** |  |  |  |
| --- | --- | --- | --- |
| DRC | 4834 | 0 | **0.0** |
| Uganda | 403 | 400 | **99.3** |
| United Arab Emirates | 743 | 389 | **52.4** |
| South Sudan | 64 | 64 | **100** |
| Kenya | 2.108 | 26 | **1.2** |
| India | 16 | 16 | **100** |
| Tanzania | 2.178 | 14 | **0.6** |
| South Africa | 162 | 2 | **1.2** |
| Angola | 36 | 1 | **2.8** |
| Zimbabwe | 72 | 0 | **0.0** |
| Other country | 6 | 4 | **66.7** |
| **Total** | **10,622** | **916** | **8.62** |

### Proportion of empty cigarette packs that were illicit

Overall, 8.6% (916) of the collected packs met at least one of the four criteria and were therefore classified as illicit (Table 1). 9.8% of packs from stationary retailers were illicit, and this was 2.2% of those collected from mobile retailers and 9.3% of those collected from garbage bins/streets. By province, the proportion of illicit cigarette packs ranged from 0.7% in Tshopo to 31.6% in Ituri. Very high-porosity provinces had the highest proportion of illicit cigarette packs of 31.6%, whilst this was 5.2% for those from high-porosity provinces, 3.5% for those from intermediate-porosity provinces and 9.4% for those from low-porosity provinces. 8.1% of the packs collected in rural areas were illicit, whilst this was 8.8% for urban areas (Table 2). 7.8% of packs from low-income areas and 10.2% of those from high-income areas were illicit. All of the illicit packs were of imported cigarettes, meaning that 15.8% of imported packs were illicit. When considering the different brands, 0% of Elite, Equateur, Master and Portsman were illicit, whilst Supermatch and Oris had the highest proportions of illicit packs, at 52.0% and 71.4% respectively.

None of the packs originating from the DRC and Zimbabwe were illicit, whilst 100% of those originating from South Sudan and India were illicit (Table 2). This was 99.2% of those from Uganda, 52.4% of those from the United Arab Emirates, 2.8% of those from Angola, 1.2% of those from South Africa, 1.2% of those from Kenya and 0.6% of those originating from Tanzania.

All of the illicit packs did not comply with tax stamp requirements and were therefore classified as having evaded cigarette tax (Table 3). Approximately 8.0% of the collected packs did not comply with written health warning requirements. Of the 9774 packs that complied with written health warning requirements, 0.3% (33) had the message *“Smoking is harmful to health”*, 31.7% (3,101) had *“Tobacco seriously harms your health”*, 73.1% (7,142) had *“Smoking is highly addictive”*, and none had the message *“Be careful, smoking kills.”* 5.6% of all collected packs did not indicate the prohibition of sale by/to minors, and 4.5% did not have information on tar and nicotine content.

**Table 3.**
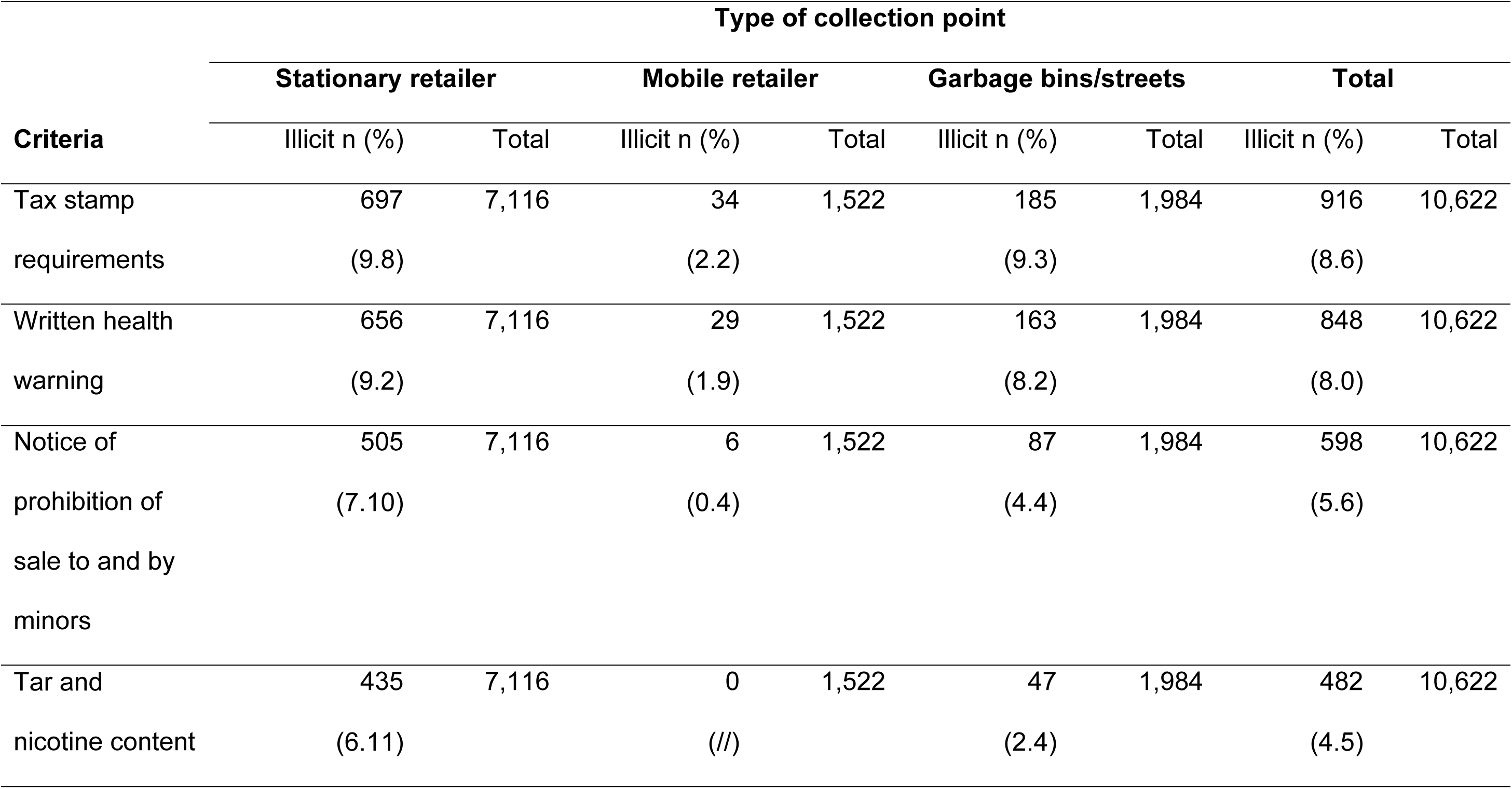
Proportion of illicit empty packs by type of collection point and illicit criteria.

When considering all imported packs with yellow stamps as illicit, 51.5% of the collected packs met at least one of the four criteria and were therefore classified as illicit. All of the illicit packs did not comply with tax stamp requirements and were therefore classified having evaded cigarette tax. The proportions that met the other criteria for illicit were similar to those found when considering all packs with yellow stamps as legal packs. The proportion of illicit cigarettes by source, province, province porosity stratum, urban/rural, area-level income group, brand, and country of origin for the sensitivity analyses are provided in S2 to S4 Tables.

### Cigarette pack retail prices

Cigarette pack retail price data were collected from 1236 retailers. The average retail price of a pack of cigarettes by brand varied from 1309 to 3185 Congolese Francs (∼US$0.50 to ∼US$1.15) (Table 4). Some brands that had a high proportion of illicit cigarette packs, such as Oris, had some of the highest average retail prices for a pack, whilst some where all packs were judged as legal, e.g., Elite, Equateur and Master, had some of the lowest prices.

**Table 4.** Average selling price of a pack of cigarettes by brand.

| Average retail price per pack of cigarettes in Congolese Francs |  |
| --- | --- |
| Brands |  |
| Oris | 3185 |
| Ambassade | 2216 |
| Stella | 2183 |
| Business | 1946 |
| Caesar | 1891 |
| Pall Mall | 1819 |
| Portsmann | 1688 |
| Monte Carlo | 1676 |
| Supermatch | 1632 |
| Master | 1596 |
| Equateur | 1554 |
| Elite | 1156 |
| Other brands | 1309 |
| **Total** | **1765** |

### Factors associated with being an illicit cigarette pack

Empty cigarette packs from low-income areas were more likely to be illicit (aOR 1.90; [95 CI: 1.48 - 2.43]) than those collected from high-income areas (Table 5). Cigarette packs sold in intermediate-porosity (aOR 3.36; [95% CI: 1.72 - 6.57]) and high-porosity (aOR 17.15; [95% CI: 7.48 - 39.31]) strata were more likely to be illicit than those in the low-porosity stratum.

**Table 5.**
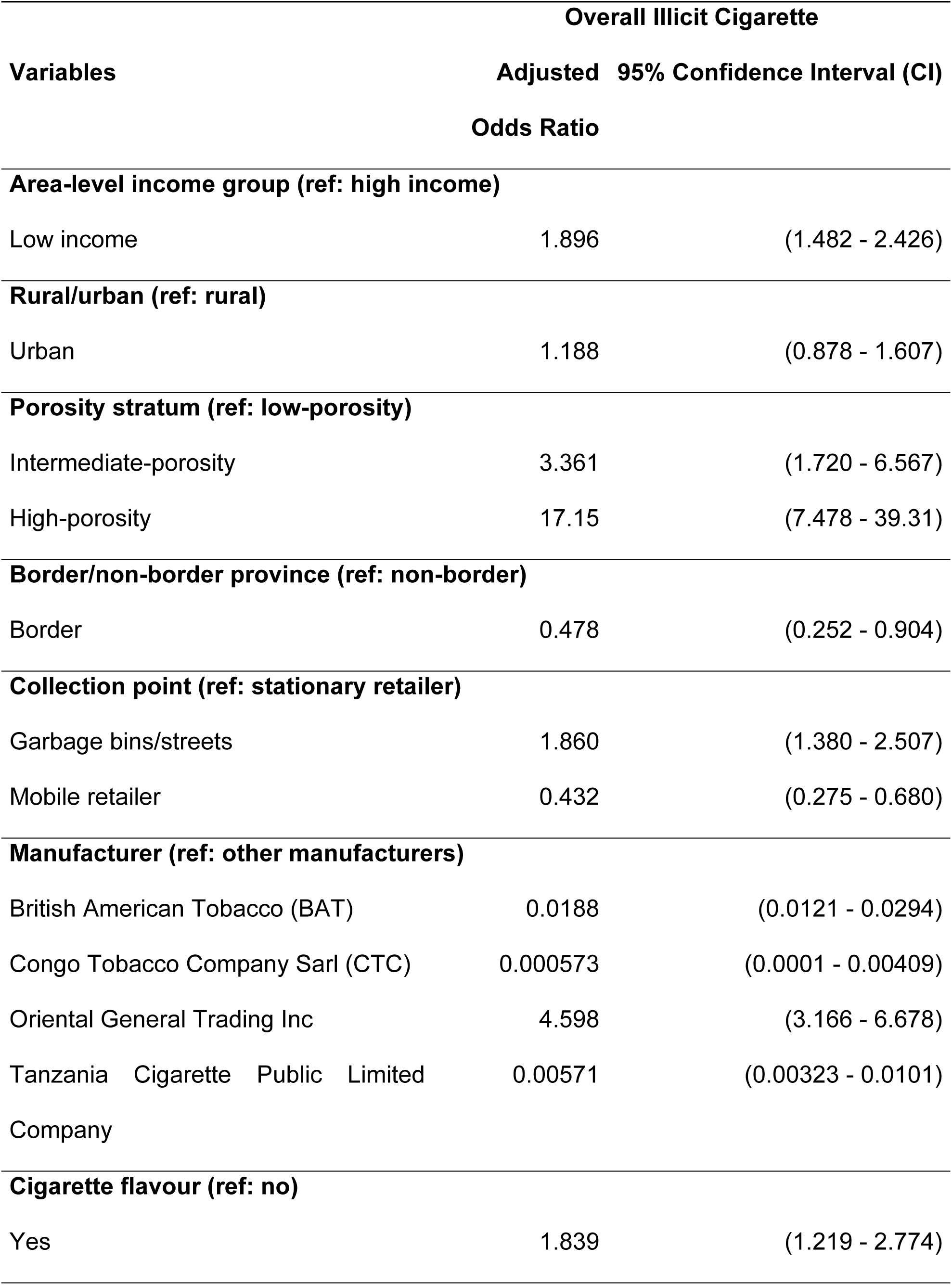
Factor associated with illicit cigarette packs.

| Variables | Overall Illicit Cigarette |  |
| --- | --- | --- |
|  | Adjusted Odds Ratio | 95% Confidence Interval (CI) |
| **Area-level income group (ref: high income)** |  |  |
| Low income | 1.896 | (1.482 - 2.426) |
| **Rural/urban (ref: rural)** |  |  |
| Urban | 1.188 | (0.878 - 1.607) |
| **Porosity stratum (ref: low-porosity)** |  |  |
| Intermediate-porosity | 3.361 | (1.720 - 6.567) |
| High-porosity | 17.15 | (7.478 - 39.31) |
| **Border/non-border province (ref: non-border)** |  |  |
| Border | 0.478 | (0.252 - 0.904) |
| **Collection point (ref: stationary retailer)** |  |  |
| Garbage bins/streets | 1.860 | (1.380 - 2.507) |
| Mobile retailer | 0.432 | (0.275 - 0.680) |
| **Manufacturer (ref: other manufacturers)** |  |  |
| British American Tobacco (BAT) | 0.0188 | (0.0121 - 0.0294) |
| Congo Tobacco Company Sarl (CTC) | 0.000573 | (0.0001 - 0.00409) |
| Oriental General Trading Inc | 4.598 | (3.166 - 6.678) |
| Tanzania Cigarette Public Limited Company | 0.00571 | (0.00323 - 0.0101) |
| **Cigarette flavour (ref: no)** |  |  |
| Yes | 1.839 | (1.219 - 2.774) |

Cigarette packs from border provinces were less likely to be illicit (aOR 0.48; [95% CI: 0.25 - 0.90]) than those from non-border provinces. Cigarette packs collected from mobile retailers were less likely to be illicit (aOR 0.43; [95% CI: 0.28 - 0.68]), while those collected from the garbage bins/streets were more likely (aOR 1.90; [95% CI: 1.38 - 2.51]) to be illicit, than those collected from the stationary retailers. Cigarette packs that were flavoured were more likely to be illicit than those that were not flavoured (aOR 1.84; [95% CI: 1.22 - 2.77]). There was no statistically significant difference between those collected from urban and rural HZs with regard to the likelihood of being illicit (aOR 1.19; [95% CI: 0.88 - 1.61]). The odds of being illicit when considering the different cigarette manufacturers are also shown in Table 5.

The relationships between each of the independent variables and three of the illicit criteria (i.e., cigarette tax evasion/non-compliance with tax stamp requirements, non-compliance with written health warning requirements, or absence of the notice indicating the prohibition of sale by/to minors) were similar to those between the each of the independent variables and overall illicit classification in both magnitude and direction (Table 6). The only differences were for 1) rural/urban HZs where packs from urban HZs were more likely not to have the notice indicating the prohibition of sale by/to minors than those from rural HZs (aOR 2.928; [95% CI: 2.022 - 4.240]); and 2) for porosity stratum where the association was stronger for high-porosity stratum (aOR 278.9; [95% CI: 81.66 - 952.5]).

**Table 6.**
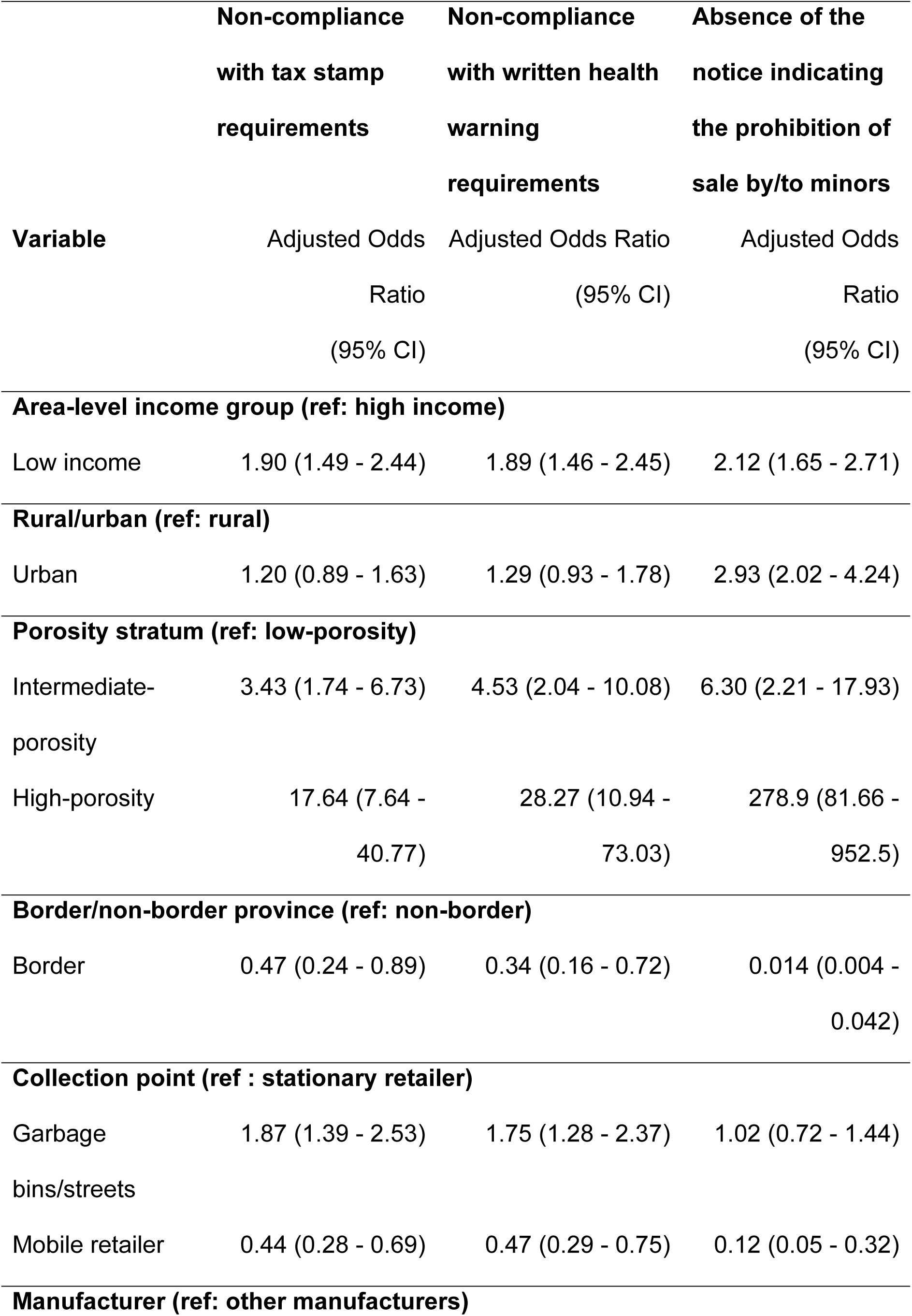
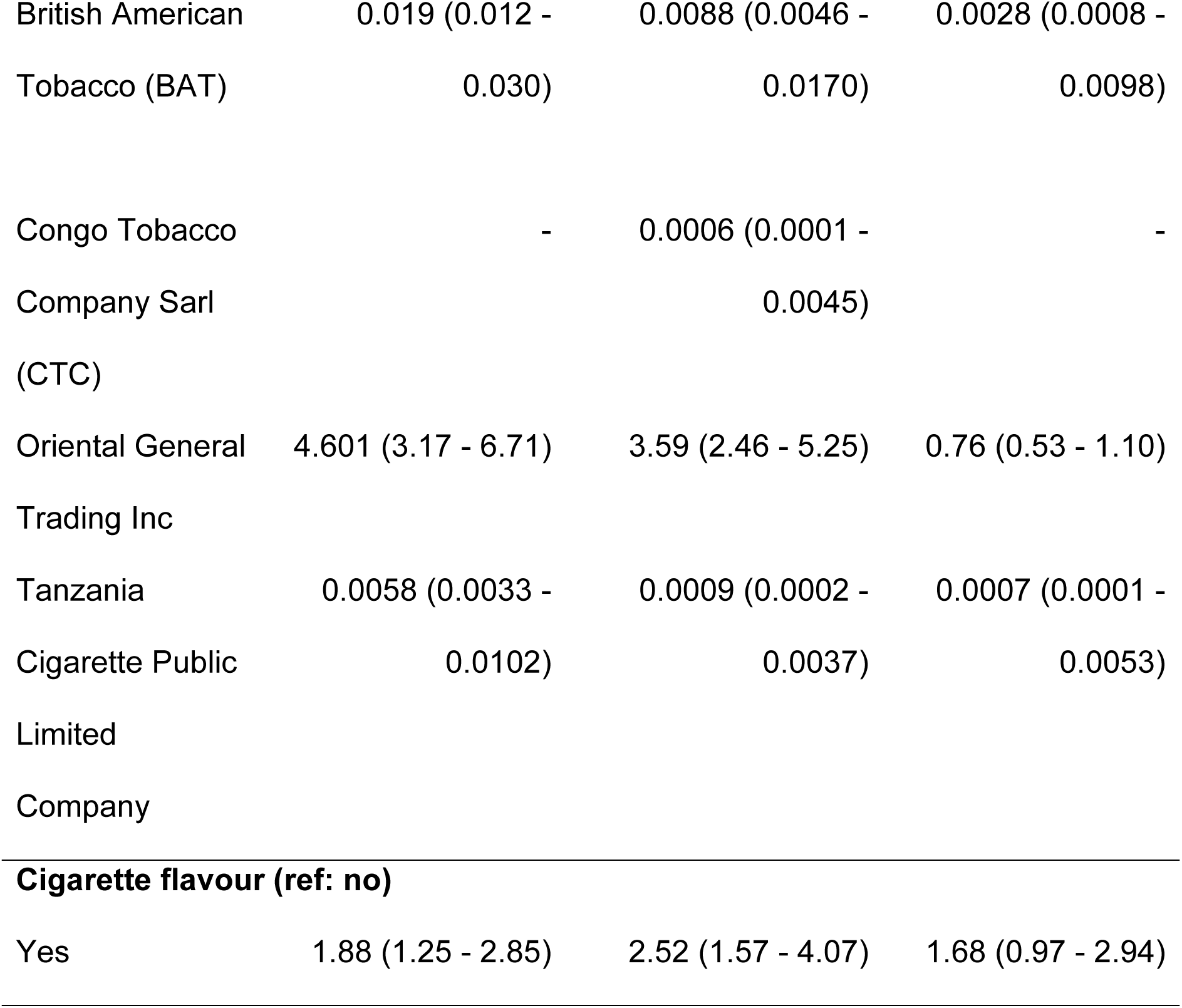
Factors associated with illicit cigarettes by criteria.

|  | **Non-compliance**<br>**with tax stamp**<br>**requirements** | **Non-compliance**<br>**with written health**<br>**warning**<br>**requirements** | **Absence of the**<br>**notice indicating**<br>**the prohibition of**<br>**sale by/to minors** |
| --- | --- | --- | --- |
| **Variable** | Adjusted Odds<br>Ratio<br>(95% CI) | Adjusted Odds Ratio<br>(95% CI) | Adjusted Odds<br>Ratio<br>(95% CI) |
| **Area-level income group (ref: high income)** |  |  |  |
| Low income | 1.90 (1.49 - 2.44) | 1.89 (1.46 - 2.45) | 2.12 (1.65 - 2.71) |
| **Rural/urban (ref: rural)** |  |  |  |
| Urban | 1.20 (0.89 - 1.63) | 1.29 (0.93 - 1.78) | 2.93 (2.02 - 4.24) |
| **Porosity stratum (ref: low-porosity)** |  |  |  |
| Intermediate-porosity | 3.43 (1.74 - 6.73) | 4.53 (2.04 - 10.08) | 6.30 (2.21 - 17.93) |
| High-porosity | 17.64 (7.64 -<br>40.77) | 28.27 (10.94 -<br>73.03) | 278.9 (81.66 -<br>952.5) |
| **Border/non-border province (ref: non-border)** |  |  |  |
| Border | 0.47 (0.24 - 0.89) | 0.34 (0.16 - 0.72) | 0.014 (0.004 -<br>0.042) |
| **Collection point (ref : stationary retailer)** |  |  |  |
| Garbage<br>bins/streets | 1.87 (1.39 - 2.53) | 1.75 (1.28 - 2.37) | 1.02 (0.72 - 1.44) |
| Mobile retailer | 0.44 (0.28 - 0.69) | 0.47 (0.29 - 0.75) | 0.12 (0.05 - 0.32) |
| **Manufacturer (ref: other manufacturers)** |  |  |  |
| British American Tobacco (BAT) | 0.019 (0.012 - 0.030) | 0.0088 (0.0046 - 0.0170) | 0.0028 (0.0008 - 0.0098) |
| Congo Tobacco Company Sarl (CTC) | - | 0.0006 (0.0001 - 0.0045) | - |
| Oriental General Trading Inc | 4.601 (3.17 - 6.71) | 3.59 (2.46 - 5.25) | 0.76 (0.53 - 1.10) |
| Tanzania Cigarette Public Limited Company | 0.0058 (0.0033 - 0.0102) | 0.0009 (0.0002 - 0.0037) | 0.0007 (0.0001 - 0.0053) |
| **Cigarette flavour (ref: no)** |  |  |  |
| Yes | 1.88 (1.25 - 2.85) | 2.52 (1.57 - 4.07) | 1.68 (0.97 - 2.94) |

## Discussion

From our primary analysis, in the DRC, illicit cigarettes constitute approximately 8.6% of the total cigarette market base. All of the illicit packs did not comply with tax stamp requirements and were therefore classified as having evaded cigarette tax. 8.0% of the collected packs did not comply with written health warning requirements. 5.6% of all collected packs did not indicate the prohibition of sale by/to minors, and 4.5% did not have information on tar and nicotine content. The chance of being an illicit cigarette pack was higher for packs collected from low-income neighbourhoods than those from high-income neighbourhoods, and increased with increasing level of province porosity (i.e., susceptibility to armed conflict/insecurity). Flavoured cigarette packs were more likely to be illicit than those for unflavoured cigarettes. Packs that were collected from garbage bins/streets were more likely, whilst those collected from mobile retailers were less likely, to be illicit than those collected from stationary retailers. Those collected from border provinces were less likely to be illicit than those collected from non-border provinces. There was no statistically significant difference in the likelihood of being an illicit pack between those collected from urban HZs and those from rural HZs.

Other Sub-Saharan African country studies have shown that illicit cigarettes constitute 8.6% of the cigarette market in the Gambia [20] and 12.2% in Zambia [21)], which is consistent with our findings. Global estimates suggest that on average, the illicit cigarette trade market share is ∼17% for low-income countries [10]. Other Sub-Saharan African countries, however, have reported higher illicit trade market shares, for example Ghana (20%) [11], Ethiopia (45.4%) [22] and South Africa (54%) [23]. This could be because, in these countries, illegal cigarettes are on average cheaper than legal purchases, whereas our results suggest an opposite trend in the DRC with illegal cigarettes, all of which were imported, being more expensive on average than legal cigarettes. In addition, contrary to findings from other Sub-Saharan African country studies [11, 22], packs that were collected from border provinces were less likely to be illicit than those collected from non-border provinces. This could suggest better enforcement and compliance monitoring in border than non-border provinces in the DRC.

We also found that the yellow stamp, an old stamp that was used for both imported and locally manufactured cigarettes, which should have been phased out in 2022, is currently still being used in the DRC. If all imported packs with the yellow stamps are considered illicit, 51.5% of the cigarette market base becomes illicit.

The majority of the packs carried the written health warning *“Smoking is highly addictive”*, whilst none had the message “*Be careful, smoking kills*.” Well-designed health warnings and messages on tobacco product packages reduce tobacco consumption by increasing public awareness of the negative health effects of tobacco use. However, the tobacco industry is known to undermine the effectiveness of health warnings in many ways, including through the use of weak message content and opposing strengthened warnings [24].

### Policy and practice implications

Our main analysis suggests that illicit cigarette trade in the DRC is low. However, there is still a need to secure the cigarette supply chain to counter the supply of illicit cigarettes in the DRC. In particular, there is a need to strengthen border controls, resolve the issue of the old yellow stamp, establish a secured track and trace system for cigarettes, and strengthen enforcement and compliance monitoring both centrally and at the provincial level. The DRC should also strengthen its political commitment to combating illicit cigarette trade by ratifying the WHO Protocol on Illicit Tobacco Trade. In addition, there is a need to strengthen health warning requirements by ensuring strong message content on all cigarette packs.

### Implications for future research

There is a need to continuously monitor illicit cigarette trade over time in the DRC in order to facilitate timely action, and enable the evaluation of the impact of any new tobacco control measures (e.g., tax and price measures), where necessary. Regular monitoring can, for example, help counter the tobacco industry’s narrative that increasing taxes result in considerable increases in illicit cigarette trade. Our study suggests that non-compliance with cigarette tax stamp requirements is a good indicator of whether a cigarette pack is legal/illicit in the DRC. This means that, if the required data are available, regular monitoring could be more efficiently achieved by measuring the difference between consumption and tax paid sales (i.e., gap analysis) [14].

Different health warnings are applicable to, and resonate with, different people [24]. It is therefore important to investigate the effectiveness of the different health warnings that are being used in the DRC to ensure that those that are used are the most effective. This should include the evaluation of their effectiveness in deterring young people from initiating smoking, and motivating those who smoke to quit.

### Strengths and limitations

This study is the first of its kind to estimate the illicit cigarette market share in the DRC. Although our study was limited to eight out of the 26 provinces of the DRC, we employed stratified, multistage random sampling to enhance the representativeness and generalizability of the findings. Our study relied on retailers providing us with empty cigarette packs, and it is possible that some retailers did not provide us with all their illegal packs, leading to underestimation of the illicit cigarette market share [11]. We tried to mitigate this by concealing our specific interest in illicit cigarette packs: at informed consent, we told retailers that we were interested in the characteristics of the packs. We also compensated retailers with 2,000 Congolese Francs (∼US$1) for the time they spent gathering the cigarette packs. These procedures were approved by the ethics committee. Our reporting followed the Strengthening the Reporting of Observational Studies in Epidemiology (STROBE) checklist for cross-sectional studies [25], and the data used in the writing of this article is publicly available [26].

### Conclusions

Our study found that 8.6% of the cigarette packs collected were illicit, and most of the illicit cigarettes were imported from other countries. There is a need to secure the cigarette supply chain to counter the supply of illicit cigarettes in the DRC. This includes strengthening border controls and enforcement and compliance monitoring, and establishing a secured track and trace system for cigarettes. Ratifying the WHO Protocol on Illicit Tobacco Trade will strengthen the DRC’s political commitment to combating illicit cigarette trade.

## Data Availability

The datasets generated and/or analysed during the current study are available in the Zenodo repository, https://doi.org/10.5281/zenodo.12359650.

## Acknowledgements

This research was carried out within the framework of the Tobacco Control Data Initiative (TCDI). TCDI is a project that encompasses six African nations (the Democratic Republic of the Congo, Ethiopia, Kenya, Nigeria, South Africa, and Zambia). Its objective is to comprehend the tobacco-related data requirements of these countries, assess existing data, pinpoint gaps in available tobacco data, gather new data to address those gaps, and create tools to assist policymakers in utilising crucial data more effectively for shaping tobacco policies. The initiative is spearheaded by Development Gateway: an IREX Venture (DG), a global nonprofit organisation specialising in data-driven development, in collaboration with the University of Cape Town’s Research Unit on the Economics of Excisable Products (REEP). We would like to express our deep gratitude to the Ministère de la Santé Publique, Hygiène et Prévoyance Sociale of the DRC and, specifically, the Programme National de Lutte Contre la Toxicomanie et les Substance Toxiques for their exceptional commitment to the realisation of this study. We also thank the cigarette retailers who collected and provided us with the cigarette packs that were analysed in this study; and the field investigators who collected them from the retailers and examined them to extract the study data. Our gratitude also goes to members of the Study Steering Committee for their guidance and advice throughout the study.

## Supporting information

**S1 Fig. Provinces by strata**

**S2 Fig. Illustration of sampling provinces, health zones and health areas by stratum**

**S1 Table. Participating health zones and health areas by province and urban/rural classification**

**S2 Table. Proportion of illicit empty packs by type of collection point, porosity and province with all imported yellow stamps as illicit**

**S3 Table. Proportion of illicit empty packs by rural/urban, area-level income group, brand and country of origin with all imported yellow stamps as illicit S4 Table. Proportion of illicit empty packs by type of collection point and illicit criteria with all imported yellow stamps as illicit.**

## REFERENCES

1. INS. Enquête par grappes à indicateurs multiples, 2017-2018, rapport de résultats de l’enquête. Kinshasa: République Démocratique du Congo; 2019.

2. World Health Organization. WHO report on the global tobacco epidemic, 2021: addressing new and emerging products. Geneva: World Health Organization; 2021.

3. United Nations Treaty Collection. Chapter IX: Health. 4. WHO Framework Convention on Tobacco Control, Geneva, 21 May 2003. https://treaties.un.org/pages/ViewDetails.aspx?src=TREATY&mtdsg_no=IX-4&chapter=9&clang=_en. Accessed 05 Jul 2024.

4. United Nations. Treaty collections: 4. a Protocol to Eliminate Illicit Trade in Tobacco Products. https://treaties.un.org/Pages/ViewDetails.aspx?src=TREATY&mtdsg_no=IX-4-a&chapter=9&clang=_en. Accessed 05 Jul 2024.

5. Guindon GE, Tobin S, Yach D. Trends and affordability of cigarette prices: ample room for tax increases and related health gains. Tob Control. 2002;11:35–43. 10.1136/tc.11.1.35.

6. Felsinger R, Groman E. Price policy and taxation as effective strategies for tobacco control. Front Public Health. 2022;10:851740. 10.3389/fpubh.2022.851740.

7. Paraje G, Stoklosa M, Blecher E. Illicit trade in tobacco products: recent trends and coming challenges. Tob Control. 2022;31:257–62. doi: 10.1136/tobaccocontrol-2021-056557.

8. Bader P, Boisclair D, Ferrence R. Effects of tobacco taxation and pricing on smoking behavior in high risk populations: a knowledge synthesis. Int J Environ Res Public Health. 2011;8:4118–39. 10.3390/ijerph8114118.

9. World Health Organization. Protocol to Eliminate Illicit Trade in Tobacco Products. 2013. https://fctc.who.int/protocol/overview. Accessed 08 Apr 2024.

10. Joossens L, Merriman D, Ross H, Raw M. The impact of eliminating the global illicit cigarette trade on health and revenue. Addiction. 2010;105:1640–9. 10.1111/j.1360-0443.2010.03018.x.

11. Singh A, Ross H, Dobbie F, Gallagher A, Kinnunen T, Logo DD, et al. Extent of illicit cigarette market from single stick sales in Ghana: findings from a cross-sectional survey. BMJ Open. 2023;13:e062476. 10.1136/bmjopen-2022-062476.

12. John RM, Ross H. Illicit cigarette sales in Indian cities: findings from a retail survey. Tob Control. 2018;27:684–8. 10.1136/tobaccocontrol-2017-053999.

13. Barker DC, Wang S, Merriman D, Crosby A, Resnick EA, Chaloupka FJ. Estimating cigarette tax avoidance and evasion: evidence from a national sample of littered packs. Tob Control. 2016;25 Suppl 1:i38–i43. 10.1136/tobaccocontrol-2016-053012.

14. Stoklosa M, Paraje G, Blecher E. A toolkit on measuring illicit trade in tobacco products. A Tobacconomics and American Cancer Society Toolkit. Chicago, IL: Tobacconomics, Health Policy Center, Institute for Health Research and Policy, University of Illinois at Chicago; 2020.

15. Dobility Inc. SurveyCTO. 2024. https://www.surveycto.com/. Accessed 08 Apr 2024.

16. Institute for Global Tobacco Control. Assessing compliance with tobacco packaging and labeling regulations. Baltimore, MD: Johns Hopkins Bloomberg School of Public Health; 2020.

17. Aziani A, Calderoni F, Dugato M. Explaining the consumption of illicit cigarettes. J Quant Criminol. 2021;37(3):751–89. 10.1007/s10940-020-09465-7.

18. van der Zee K, Vellios N, van Walbeek C, Ross H. The illicit cigarette market in six South African townships. Tobacco Control. 2020;29 Suppl 4:s267. 10.1136/tobaccocontrol-2019-055136.

19. StataCorp LLC. Stata. 2024. https://www.stata.com/. Accessed 08 Apr 2024.

20. Chisha Z, Janneh ML, Ross H. Consumption of legal and illegal cigarettes in the Gambia. Tob Control. 2020;29 Suppl 4:s254–s9. 10.1136/tobaccocontrol-2019-055055.

21. Zyambo C, Phiri MM, Mwamulela W, Zulu R, Maka M, Camara A, et al. Illicit cigarette trade and tax evasion in Zambia: findings from the Tobacco Control Data Initiative 2023. BMJ Open. 2024.

22. Dauchy E, Ross H. Is illicit cigarette market a threat to tobacco control in Ethiopia? Nicotine Tob Res. 2022;24:1228–33. 10.1093/ntr/ntac021.

23. Vellios N, van Walbeek C, Ross H. Illicit cigarette trade in South Africa: 2002-2017. Tob Control. 2020;29 Suppl 4:s234–s42. 10.1136/tobaccocontrol-2018-054798.

24. Cunningham R. Tobacco package health warnings: a global success story. Tob Control. 2022;31:272–283. 10.1136/tobaccocontrol-2021-056560.

25. von Elm E, Altman DG, Egger M, Pocock SJ, Gotzsche PC, Vandenbroucke JP. The Strengthening the Reporting of Observational Studies in Epidemiology (STROBE) Statement: guidelines for reporting observational studies. J Clin Epidemiol. 2008;61:344–349. 10.1016/j.jclinepi.2007.11.008.

26. Development Gateway: An IREX Venture. The illicit cigarette market in the DRC: datasets from a cross-sectional study of empty cigarette packs [Data set]. Zenodo. 2024. 10.5281/zenodo.12359650.

